# Funding and COVID-19 Research Priorities - Are the research needs for Africa being met?

**DOI:** 10.1101/2020.10.12.20211565

**Authors:** Emilia Antonio, Moses Alobo, Marta Tufet Bayona, Kevin Marsh, Alice Norton

## Abstract

**Introduction:** Emerging data from Africa indicates remarkably low numbers of reported COVID-19 deaths despite high levels of disease transmission. However evolution of these trends as the pandemic progresses remains unknown. More certain are the devastating long-term impacts of the pandemic on health and development evident globally. Research tailored to the unique needs of African countries is crucial.

UKCDR and GloPID-R have launched a tracker of funded COVID-19 projects mapped to the WHO research priorities and research priorities of Africa and less-resourced countries and published a baseline analysis of a Living Systematic Review (LSR) of these projects.

**Methods:** In-depth analyses of the baseline LSR for COVID-19 funded research projects in Africa (as of 15^th^ July 2020) to determine the funding landscape and alignment of the projects to research priorities of relevance to Africa.

**Results:** The limited COVID-19 related research across Africa appears to be supported mainly by international funding, especially from Europe, although with notably limited funding from United States-based funders. At the time of this analysis no research projects funded by an African-based funder were identified in the tracker although there are several active funding calls geared at research in Africa and there may be funding data which has not been made publicly available.

Many projects mapped to the WHO research priorities and 5 particular gaps in research funding were identified namely: investigating the role of children in COVID-19 transmission; effective modes of community engagement; health systems research; communication of uncertainties surrounding mother-to-child transmission of COVID-19; and identifying ways to promote international cooperation. Capacity strengthening was identified as a dominant theme in funded research project plans.

**Conclusions:** We found significantly lower funding investments in COVID-19 research in Africa compared to High-Income Countries, seven months into the pandemic, indicating a paucity of research targeting the research priorities of relevance to Africa.

**Summary Box:** 

**What is already known?:**

- There has been a swift global research response to the COVID-19 pandemic guided by priorities outlined in the WHO Research Roadmap and hundreds of research activities have rapidly been commissioned.
- The research priorities for Africa are likely to be influenced by unique contextual factors which could worsen the prognosis of infections and influence measures for disease prevention and control and indirect long-term disease impacts.
- Remarkably, there has been a low number of reported COVID-19 mortalities despite emerging evidence of high levels of transmission in Africa.

**What are the new findings?:**

- We present the most comprehensive assessment of COVID-19 research investments in Africa seven months into this pandemic and found significantly less research investments in Africa, given that only 84 out of 1858 research projects identified globally involved at least one African country.
- Several important gaps in funded research in Africa were identified indicating some areas requiring greater research focus.
- The dominant capacity strengthening theme in funded research projects highlights insufficient pandemic research preparedness of African countries.

**What do the new findings imply?:**

- An assessment of the alignment of funded research projects in Africa to important global and regional research priorities is imperative for gaining key insights into the trends of disease, guiding research funding investments, prevention and control strategies and learning lessons for future pandemics.
- In this context of limited resources, investments in research in Africa must be targeted at the most pressing research needs for effective control of this pandemic.

## Introduction

### Pandemic preparedness

The Coronavirus 2019 (COVID-19) pandemic hit at a time when pandemic preparedness was at the fore of global health policy but under-resourced. The 2014-2016 West Africa Ebola outbreak had exposed glaring gaps in the international outbreak response mechanism and, in its wake, many evaluation panels were commissioned to consider lessons learnt for response to future outbreaks. Several global and regional initiatives were commissioned to support the activities of existing initiatives such as Global Outbreak Alert and Response Network (GOARN) and African Coalition for Epidemic Research, Response and Training (ALERRT). These include Regional Disease Surveillance Systems Enhancement (REDISSE) for strengthening disease surveillance in West Africa, Coalition for Epidemic Preparedness and Innovation (CEPI) and a World Bank-funded $500 million bonds scheme to promote pandemic preparedness in developing countries (1) (2). Importantly, the Africa Centres for Diseases Control, a Pan African initiative to promote collaboration and partnership among African nations and advance public health was established. The WHO R&D Blueprint was also launched, highlighting priority pathogens of outbreak potential and developing a coordinated research response mechanism in preparation for disease outbreaks (3).

Despite these laudable initiatives, Joint External Evaluation (JEE) scores, representing a voluntary evaluation of country-level preparedness benchmarks outlined in the International Health Regulations (IHR), were strikingly lower across Africa and for lower-income countries in general in 2019 indicating a lack of pandemic preparedness (4)(5). These findings resonated with global preparedness levels outlined in the maiden report of the Global Preparedness Monitoring Board (GPMB), in September 2019 just 2 months prior to the onset of the COVID-19 pandemic. The report took cognisance of recommendations from the 2009 H1N1 pandemic and the 2014-2016 Ebola outbreak and concluded global pandemic preparedness was inadequate. Further, it set out the crucial steps to be taken by governments, donors and funders to ensure sustainable preparedness plans in response to the next pandemic (6).

### Covid-19 In Africa

As of 30^th^ September 2020, there were 1,182,927confirmed COVID-19 infections and 25,881 deaths in Africa, representing 3.5 % and 2.6 % of global infections and mortalities respectively (7). Despite emerging evidence of high levels of transmission of the virus, Africa is one of the least directly impacted continents when disease burden alone is considered and there is keen interest in the evolution of these trends as the pandemic progresses. Emerging evidence from Europe and the United States are indicative of severe long-term sequelae following even mild COVID-19 infections (8) (9) (10). These aftereffects, the magnitude of which is yet to be determined, could further burden health systems in Africa. In spite of the apparently low direct mortality, the cumulative effects of comprehensive control efforts are projected to have major long-term impacts which could potentially offset decades of health, economic and developmental gains in the sub-region.

Africa is made up of diverse countries with unique contextual characteristics likely to influence COVID-19 outcomes, prevention, control and management. The projected transgenerational impacts of suspended education, immunizations and maternal and child health programmes resulting from disruptions caused by the pandemic are grave. The observation of a higher proportion of deaths among younger people living with HIV in South Africa speaks to the influence of infectious diseases on COVID-19 outcomes (11). Importantly, tuberculosis, malaria and other infectious disease burdens which are disproportionately higher in Africa could potentially worsen the prognosis of COVID-19 infections.

Coupled with the aforementioned, the rising burden of non-communicable diseases has stretched existing health systems to capacity, and COVID-19 could rapidly overwhelm health systems, as has been witnessed across the globe in even the best resourced countries. Overcrowded informal settlements and refugee camps, inadequate water, sanitation and hygiene (WASH) and high illiteracy levels, which hinder understanding of diseases and fuel misinformation, may make compliance with public health interventions for COVID-19 control difficult in some settings. The interplay of these among many factors gives rise to multiple vulnerabilities which are likely to influence the impact of COVID-19 in Africa.

### Research Priorities

The global research response to COVID-19 has been governed by the WHO’s *Coordinated Global Roadmap: 2019 Novel Coronavirus* in line with the WHO R&D Blueprint mechanism, which was rapidly mobilised at the onset of the outbreak (12). This Roadmap outlines 9 mid-to long-term broad research priority actions and corresponding sub-priorities for controlling the pandemic. Following the declaration of a global pandemic, the African Academy of Sciences (AAS) engaged African researchers through a survey and consultative workshop to assess the applicability of these research priorities to Africa and found a general agreement of African researchers with the WHO research priorities. However, important context-relevant research priorities falling outside the WHO framework were also identified and outlined in the *Research and Development goals for COVID-19 in Africa Report* (13).

In May 2020, a further collaborative effort between the United Kingdom Collaborative on Development Research (UKCDR), AAS and the Global Health Network (TGHN) led to a mixed methods study to determine the R&D priorities for COVID-19 by building on both the WHO Roadmap and the prior AAS study, with a special focus on less-resourced countries. This study found several WHO research priorities which required greater research emphasis and, more importantly, outlined new research priority areas which were not captured in either the WHO framework or the AAS study (14).

### Study Aim

In order to guide funding investments in research and promptly identify gaps and synergies to maximise the impact of research for this and future pandemics, UKCDR and Global Research Collaboration for Infectious Diseases Preparedness (GloPID-R) have launched the *COVID CIRCLE* (15). This learning and coordination initiative has, as part of its activities, initiated a live COVID-19 Research Project Tracker, which identifies research by key global funders classified against the WHO research priorities (16). A section of the Tracker is dedicated to monitoring the WHO International Clinical Trials Registry Platform (ICTRP) for COVID-19 clinical trials and an analysis of clinical trials involving Official Development Assistance (ODA) recipient countries is publicly available in the Tracker. Further, a living systematic review (LSR) of funded research projects has been started and the Baseline results and study protocol were published in Wellcome Open Access in September 2020 (17).

Building on this review, this article offers an in-depth analysis of the funded COVID-19 research projects in Africa, presenting the most comprehensive overview of funded research activities in Africa to date (to the best of our knowledge).

## Methods

In-depth analyses of findings from the LSR previously mentioned was done to determine the state of funded research in Africa. Similar methodology to the LSR were employed where both descriptive and thematic analyses were done using *Microsoft Excel 2019*. Key variables extracted for the LSR and study protocol are available at Wellcome Open Research (17). All projects listed in the UKCDR/GloPID-R Tracker (the Tracker), as of 15th July 2020 were eligible.

### Mapping to Research priorities

Research projects were mapped to the nine WHO broad research priory areas and corresponding sub-priorities and research priorities for Africa and less-resourced countries (14). The detailed methodology for data coding onto the Tracker is outlined in the LSR protocol.

Data entry was carried out cooperatively in a nine-person team and verification done by an independent reviewer to ensure consistency across extracted data. Projects were first assessed for a primary WHO research priority area(s) of focus. ‘N/A’ was assigned for projects which focused on innovation, research implementation/administration or clearly fell outside the WHO priorities. Next, projects were assigned to a WHO sub-priority area(s) of research focus. This process was repeated to assign broad secondary research priority and sub-priority area(s) of focus where indicated. Hence, all projects were assigned to multiple primary and/or secondary research priority areas of research focus where possible.

### Geographical distribution, funders and funding amounts

Subsequently, research projects were stratified by continent and all projects involving at least one African country (as defined by the African Union) were included in the analysis. Descriptive analyses of funding amounts, funders and research locations were made. Further analyses for potential gaps in research funding and thematic analyses of projects involving capacity strengthening were done.

### Comparative analysis

Research locations and funding investments were compared between two sections of the Tracker (funded research projects and *WHO ICTRP Analysis of Trials in DAC List Countries*, last updated 20^th^ June 2020 and G-Finder COVID-19 R&D Tracker, last updated on 18^th^ September 2020 and accessed on 20^th^ August 2020 and 29^th^ September 2020 respectively.

## Results

### Funding landscape of Research in Africa

Eighty-four projects of the 1,858 included in the LSR involved at least one African country. Four projects with non-specific country details but listed as being conducted in ‘multiple African countries’ or in ‘Africa’ were included in the analysis. Thirty-six African countries were represented in the Tracker and more West-African countries than countries from the other sub-regions were involved in research as shown in Figure 1. The majority of research projects involved Uganda (15 projects), followed by Burkina Faso and South Africa (11 projects each) and Kenya (10 projects). The paucity of research involving African countries is consistent across the Tracker’s sections and G-finder COVID-19 R&D Tracker with only minor differences. Whereas the Funded Research Project Tracker found a dominance of research projects in Uganda, the *WHO ICTRP: Analysis of Trials in DAC List Countries*, found 111 research projects with Egypt alone involved in 72 clinical trials, mostly primarily sponsored by local universities, as of 20^th^ June 2020. Similarly, *G-finder COVID-19 R&D Tracker* lists few R&D projects under research in Africa (18). Least Developed Countries (LDCs) which included 33 of the 54 countries dominated. There are 18 countries with no documented research projects in the Tracker. Twelve funders identified in the Tracker fund COVID-19 research in Africa and of these French Agency for Research on AIDS and Viral Hepatitis (ANRS), European & Developing Countries Clinical Trials Partnership (EDCTP) and UK Research and Innovation (UKRI) funded the most projects. Many projects are carried out across multiple countries as shown in Figure 2. Interestingly, UKRI funds many research projects in Gambia and Uganda, the locations of the Medical Research Council units, demonstrating the benefits of long-term investments and research links. Some funders funded projects which studied COVID-19 in existing research cohorts in African countries. Six such projects were identified and this likely highlights the usefulness of existing research networks, which can be rapidly pivoted, in emergency response to outbreaks.

**Figure 1:**
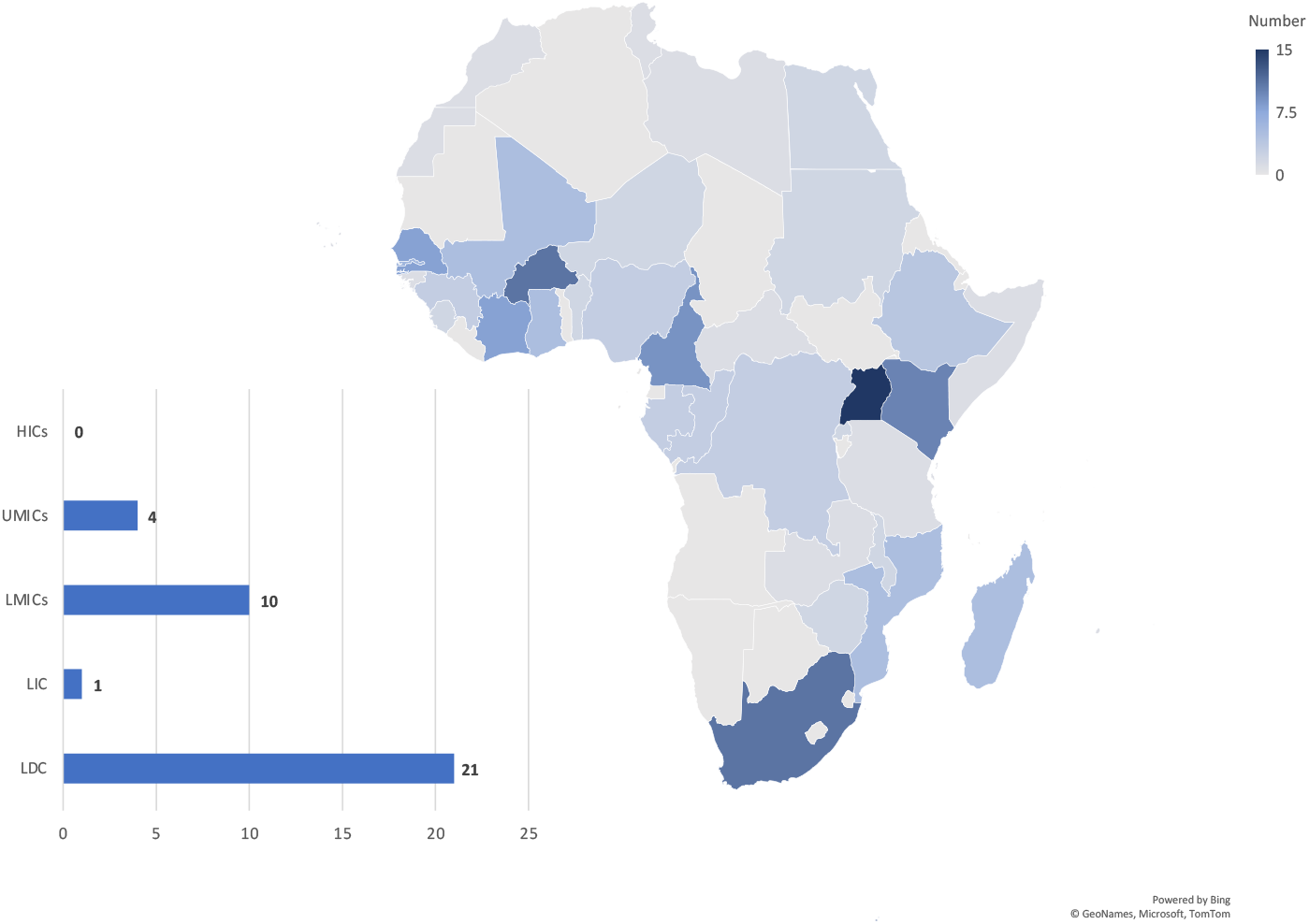
Location of COVID-19 Research Projects in Africa by Country and OECD-DAC Categories.

**Figure 2:**
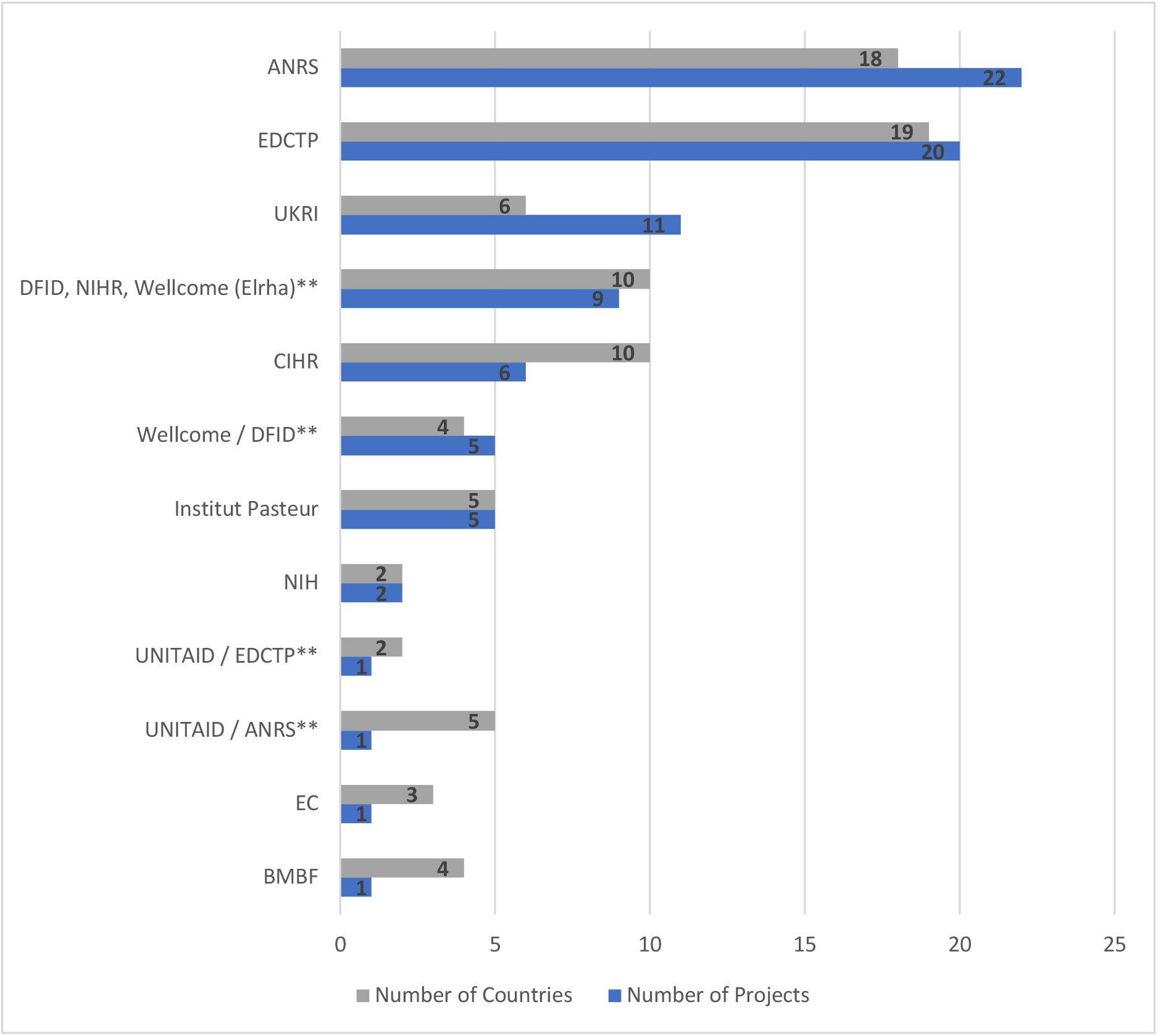
Number of Projects by Research Funder and Number of African countries across which Projects take place. *Note: ** Co-funded projects which are counted separately from other instances where funder(s) appear* Abbreviations and acronyms: **ANRS** - French Agency for Research on AIDS and Viral Hepatitis; **BMBF** - Federal Ministry of Education and Research (Germany); **CIHR** - Canadian Institutes of Health Research; **DFID** - Department for International Development (UK); **EC** - European Commission; **EDCTP** - European & Developing Countries Clinical Trials Partnership; **NIH** - National Institutes of Health (USA); **NIHR** - National Institute for Health Research; **UKRI** - UK Research and Innovation.

About $22 million of $726 million invested globally has been invested in research projects involving African countries but this value is underestimated given that only 32% of projects involving Africa included information on funding amounts. Importantly, funding amounts for EDCTP and ANRS, which are the top funders of projects involving African countries, were not available at the time of this analyses. Funding information for EDCTP projects are now available and subsequent updates of this analysis will incorporate these and any further updates on research investments made to the Tracker.

### Classification of Projects against Research priorities

#### WHO Research Priorities

Figure 3 shows most projects were classified under ‘epidemiological studies’, ‘social sciences in the outbreak response’ and ‘virus: natural history, transmission and diagnostics’. ‘Ethical considerations for research’ was the research focus of only one project. Both ‘candidate vaccine R&D’ and ‘candidate therapeutics R&D’ were the focus of few research projects in Africa; and for one project there was insufficient information to classify under a WHO priority area.

**Figure 3:**
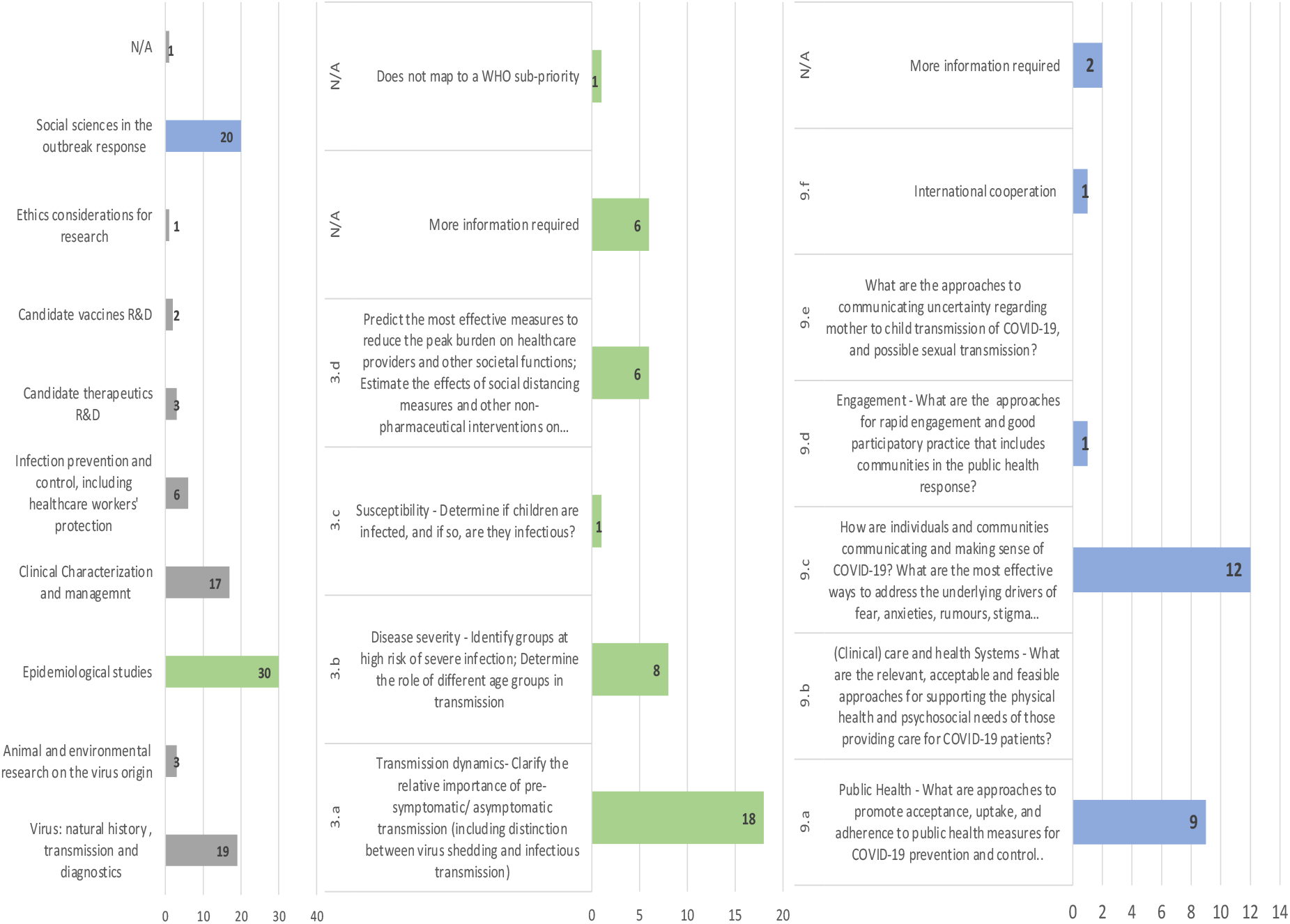
Research projects in Africa classified against the WHO Research Priorities with detailed classification of projects falling under ‘epidemiological studies’ and ‘social sciences in the outbreak response’. Bar charts show primary area of research focus only. From left to right - 1. Research projects in Africa classified against 9 WHO broad priorities; 2. Research projects classified under ‘Epidemiological studies’; 3. Research projects Classified under ‘social sciences in the outbreak response’ Note: Some projects assigned to multiple priority areas

Given that many of the priorities that emerged from the AAS and UKCDR/AAS/TGHN study fell under ‘epidemiological studies’ and ‘social sciences in the outbreak response’ projects categorised under these were analysed for gaps in research funding. The analysis revealed only one research project each focussed on disease transmission and susceptibility in children, international cooperation and feasible ways of public engagement whilst none of the projects involved health systems research or communication of uncertainties concerning COVID-19 infections and pregnancy. These findings are shown in Figure 3.

#### Research Priorities for Africa and Low-resourced countries

Few projects mapped to the ‘existing WHO research priorities requiring greater emphasis’ and priorities of Africa and less-resourced countries. Most projects that did involve understanding COVID-19 among vulnerable populations including refugees and migrants, employing technology in the pandemic response, focusing on persons living with HIV, sickle cell disease and tuberculosis and strengthening local capacity for viral genotyping as indicated in Figures 4 and 5. Capacity strengthening was a predominant theme which emerged from reviewing research projects being carried out in Africa. Of the 17 projects identified, most involved laboratory capacity strengthening activities whilst the remainder involved capacity for clinical management and surveillance.

**Figure 4:**
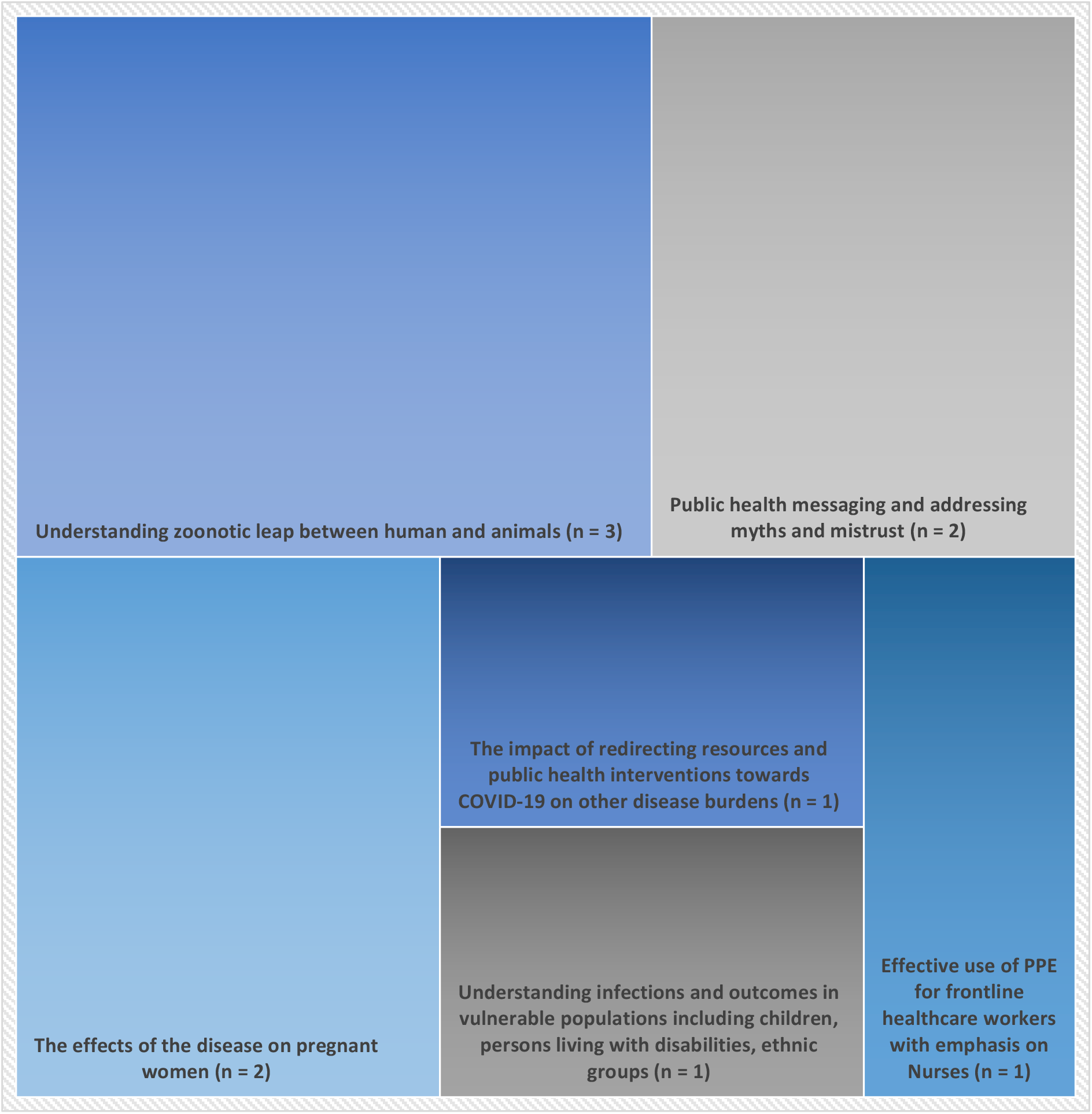
Research projects classified under ‘Existing WHO priorities requiring greater research emphasis’.

**Figure 5:**
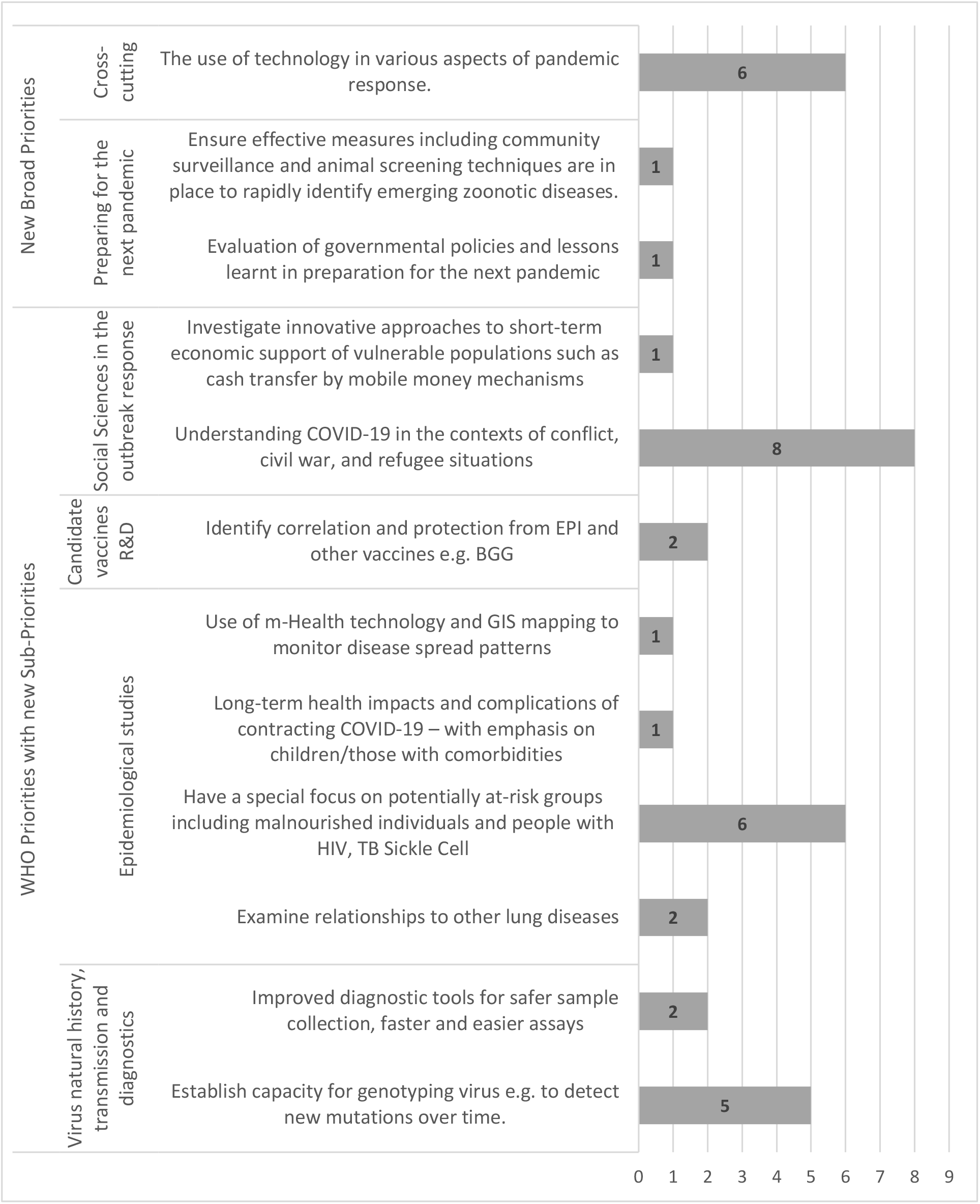
Research projects classified under the new Research priorities of Less-resourced Countries.

## Discussion

### Funding landscape of Research in Africa

In the Tracker version analysed (15^th^ July 2020), there were 1,858 COVID-19 research projects underway globally in 102 countries with only 84 (4.5%) of these projects involving at least one of 36 African countries. This finding is likely representative of the dearth of research projects in Africa given that similar findings were identified in the comparative analyses.

About 3% of total research funding ($ 22 million of $726 million spent globally) was invested in COVID-19 research in Africa representing a minute fraction of total investments by funders. Funders from Europe fund the most projects in Africa. Conspicuously underrepresented are United States-based funders which together with European funders have historically been key players in R&D funding for disease outbreaks in Africa, the most notable being the 2014 - 2016 Ebola outbreaks (19). No research projects funded by the NSF in Africa were captured in the Tracker and only 2 are funded by the NIH involving Tanzania and Madagascar. However, this trend may be shifting since Central, Eastern and Western Africa sites have been included in 11 NIH-funded grants, announced in August 2020, dedicated to the establishment of centres for research into emerging infectious diseases (20).

Also underrepresented in the funding landscape are Africa-based research funders and projects commissioned by individual country governments. Although this analysis did not capture any projects funded by Africa-based funders or governments, a review of *G-finder COVID-19 R&D Tracker: Public, philanthropic & industry funding for COVID-19 R&D* also found only a few state-funded projects in Namibia, South Africa, Nigeria and Ethiopia (18). It may well be that data on these investments are yet to be captured in either tracker or have not been made available by country governments and, this poses a significant challenge to tracking the COVID-19 research activities in Africa. Further, there are several pending research funding calls for Africa, including calls from the AAS, which might alter the findings of this analysis as the pandemic evolves.

The novelty of COVID-19 and the global scale of infections has presented unique challenges to research funders in balancing donor countries’ needs, where the pandemic peaked early, with funding of projects in less-resourced countries. Funders’ prioritization of the former may explain the trends in underfunding in Africa. Conversely, significant global funding investments have been made into diagnostics, vaccines and therapeutics to enhance disease detection and control of infections which will potentially have long-term wider benefit. Given that this is a global pandemic, it is not necessarily surprising that preliminary work in these fields tends to be most easily carried out in the best resourced settings. Indeed, a similar finding was noted after the 2014 - 2016 Ebola outbreak where the majority of R&D investments were in preclinical research in research institutions in Europe and USA (19), however context specific research is vital. The launching of the WHO access to COVID-19 tools (ACT) initiative is intended to enhance equitable and fair access to research discoveries to promote global recovery from COVID-19 moving forward (21). Seven months into this pandemic, these trends of a slow research funding response may be early indicators of the limited uptake of the GPMB recommendations for pandemic preparedness and the lack of full operationalisation of many post-Ebola initiatives, many of which had still not reached their financial targets prior to this pandemic (1).

#### Funding for COVID-19 research priorities

All projects with sufficient information for classification mapped to WHO research priorities and this signifies the alignment of researchers and funders to these priorities. There is a global lack in research projects focused on ‘Ethics considerations for research’ since this broad priority, as framed in the WHO Roadmap, outlines actions to be taken by the WHO itself including the crafting of guidance protocols for ethical research practice during the pandemic. No preclinical research projects were identified in Africa, supporting the earlier discussion concerning lack of research capacity with few clinical trials (mainly funded by EDCTP).

Further gaps in research funding for ‘epidemiological studies’ and ‘social sciences in the outbreak response’ were identified through this analysis. One project under ‘epidemiological studies’ clearly fell outside the WHO sub priorities and the new research priorities for Africa and less-resourced countries. This interesting project’s primary focus is devising innovative surveillance tools for COVID-19 mortalities in resource-limited contexts. There is a potential gap in research funding for projects to determine the role of children in COVID-19 transmission in Africa. Children, particularly those without co-morbidities, experience milder and often asymptomatic infections and their exact role in disease transmission remains unclear (22) (23). Over 85% of children born with sickle cell disease are in Africa and the high prevalence of malnutrition, HIV and tuberculosis may further worsen the prognosis of paediatric COVID-19 infections (24) (25). Further, it is challenging to distinguish some of the symptoms of COVID-19 from endemic infections such as malaria and other febrile illnesses. This challenge is likely to exacerbate under-testing of children in these settings and potentially worsen the spread of COVID-19.

Important gaps were identified for ‘social sciences in the outbreak response’ in funding for health systems research, research into effective modes for community engagement, communication of uncertainties related to COVID-19 in pregnancy and international cooperation. Given the massive shortages of health care personnel and limited health infrastructure in many African countries, health systems research during the pandemic is a crucial field of research. There have already been incidents of striking health workers in protest of insufficient personal protective equipment and support from country health ministries (26) (27) (28).

Effective communication has been an major challenge in relaying information on COVID-19, as our knowledge base continues to expand. This challenge is particularly magnified when communication of risks of mother-to-child transmission, impacts of COVID-19 on pregnancy and severity of neonatal infections, where there remain many unknowns, is considered (29) (30). The unproven risks of COVID-19 transmission in breastmilk, have to be well communicated. Breastfeeding is a well-established practice which prevents malnutrition and infectious diseases in thousands of children across Africa and the current WHO guidance favours continued breastfeeding of children with COVID-19 positive mothers as the benefits far outweigh the risks (31)(32). However, miscommunication could have negative implications for child health and survival and thus, priority should be given to research for determining the optimal approaches to engage families and communities to prevent undesirable child health outcomes.

The relevance of community engagement in Africa cannot be overemphasised as it is pivotal for understanding of and adherence to public health interventions to control COVID-19. Likewise fostering international cooperation and investigating modes of facilitating cooperation among various actors through transdisciplinary science and data sharing is crucial for control of this pandemic since “No one is safe until everyone is safe” (33).

In general, few projects mapped specifically to the additional research priorities of relevance to Africa and less-resourced countries. This finding is likely due to the overall limited research activity captured in Africa. The dominant projects identified concerned understanding COVID-19 in populations that are particularly vulnerable to adverse outcomes such as refugees and migrants, minority groups, persons with HIV and tuberculosis. Some projects fell under crosscutting uses of technology in the pandemic response and building capacity for viral genotyping. These may represent research funding gaps. However, considering only a few research projects are being conducted or funded in Africa, a more comprehensive assessment can be made once more funding calls are announced. The AAS funding call, Global Effort on COVID-19 (GECO) health research and similar funding calls, which specifically focus on low-and-middle-income-countries (LMICs), will be important to consider (34).

As several R&D candidates advance to large trials in diverse populations, inadequate research capacity can delay initiation of vital research and in the long run hinder the global research response. Laboratory capacity is particularly indispensable for the monitoring of trends in infection and determining recovery rates. It plays a key role in surveillance and clinical management which were also identified for capacity strengthening in this study. One concern is that capacity strengthening activities highlighted in this analysis may turn out to be short-lived due to their rapid mobilisation in response to the pandemic. Effective research capacity strengthening involves sustained deliberate actions geared at various aspects of the research process and at various levels of coordination at global, regional and national levels. These processes will enable countries with the greatest need to fairly and openly compete for research without compromising on quality (35). Moving forward, cooperation among research funders, enhanced mobilisation of domestic funding for capacity strengthening and periodic evaluations to guide future investments, as highlighted by the ESSENCE group of funders, are key steps for building sustained research capacity in Africa (36).

## Limitations

This analysis was based on the earlier Baseline LSR of funded COVID-19 projects initiated by the COVID CIRCLE and is similarly limited by variable completeness of data provided for classification of research projects and data on funding amounts invested in research funding. The Tracker also does not present a complete picture of repurposed research grants for COVID-19, as these details have not yet been provided by funders or are yet to be identified. There are pending funding calls related to Africa which could alter the findings of this analysis and thus this analysis can be viewed as a baseline assessment of research funding in Africa for which follow up analyses can be done to observe trends. Comparisons made to past Ebola outbreaks are made cautiously with full cognisance of the fact that this is an ongoing pandemic likely to evolve whereas findings from past outbreaks have been gleaned from retrospective review in the recovery phase.

## Conclusions

Seven months into this pandemic, this review of funded research projects in Africa has demonstrated limited funding investments by both local funders and governments, and the traditional donors and funders from previous outbreaks in Africa. The notable example here is the United States-based NIH which was a dominant donor in the Ebola outbreak of 2014 - 2016. Significant gaps in funded projects were identified in researching the role of children in COVID-19 transmission, communication of uncertainties related to antenatal and peripartum COVID-19 infections, research for feasible modes of community engagement and international cooperation and health systems research in Africa.

Few research projects mapped to the research priorities of importance to African researchers and the priorities of less-resources countries and, though this could indicate a potential gap in research funding, a more accurate assessment can be made once further funding calls for research in Africa are announced. These will be incorporated in future iterations of the Tracker LSR.

Poor research capacity and inadequate preparedness for this pandemic were demonstrated by the finding that many research projects included a capacity strengthening component. This finding may be an early indicator of limited uptake of recommendations by the GPMB Report published in 2019.

## Data Availability

All data referred to in this manuscript are from open access research platforms.
The UKCDR/GloPID-R tracker of funded COVID-19 Research Projects is available at https://www.ukcdr.org.uk/funding-landscape/covid-19-research-project-tracker/ 
Baseline Living Systematic Review of COVID-19 Funded Research Projects is available at https://wellcomeopenresearch.org/articles/5-209

https://wellcomeopenresearch.org/articles/5-209

## Acknowledgements

We are grateful to all the funders who have provided data for the Tracker to date. We also thank Nicole Advani, Adrian Bucher, Emma Clegg, Alice Cross, Henrike Grund, Sheila Mburu and Laura Scott from UKCDR and Gail Carson, Morgan Lay and Genevieve Boily-Larouche from GloPID-R for their support in the development of and coding on the Tracker.

## References

1. Ravi SJ, Snyder MR, Rivers C. Review of international efforts to strengthen the global outbreak response system since the 2014-16 West Africa Ebola Epidemic. Health Policy Plan. 2019;34(1):47–54.

2. The World Bank. World Bank Launches First-Ever Pandemic Bonds to Support $500 Million Pandemic Emergency Financing Facility [Internet]. Webpage. 2017 [cited 2020 May 31]. Available from: https://www.worldbank.org/en/news/press-release/2017/06/28/world-bank-launches-first-ever-pandemic-bonds-to-support-500-million-pandemic-emergency-financing-facility

3. World Health Organization. A research and development Blueprint for action to prevent epidemics - Plan Of Action May 2016. 2016;(May):44. Available from: https://www.who.int/blueprint/about/r_d_blueprint_plan_of_action.pdf

4. Talisuna A, Yahaya AA, Rajatonirina SC, Stephen M, Oke A, Mpairwe A, et al. Joint external evaluation of the International Health Regulation (2005) capacities: Current status and lessons learnt in the WHO African region. BMJ Glob Heal [Internet]. 2019 Nov 1 [cited 2020 Aug 19];4(6):1312. Available from: http://gh.bmj.com/

5. World Bank. Pandemic Preparedness Financing Pandemic Preparedness Financing STATUS UPDATE. 2019.

6. World Bank Group for the Global Preparedness Monitoring Board. Pandemic preparedness financing. 2019;7(7):136.

7. WHO Coronavirus Disease (COVID-19) Dashboard | WHO Coronavirus Disease (COVID-19) Dashboard [Internet]. Who. 2020 [cited 2020 Aug 19]. Available from: https://covid19.who.int/?gclid=EAIaIQobChMIiY6Ou7yn6wIVA-vtCh0xFQT8EAAYASAAEgK9JvD_BwE

8. Rubino F, Amiel SA, Zimmet P, Alberti G, Bornstein S, Eckel RH, et al. New-Onset Diabetes in Covid-19 [Internet]. Vol. 383, New England Journal of Medicine. Massachussetts Medical Society; 2020 [cited 2020 Sep 28]. p. 789–91. Available from: http://www.nejm.org/doi/10.1056/NEJMc2018688

9. Paterson RW, Brown RL, Benjamin L, Nortley R, Wiethoff S, Bharucha T, et al. The emerging spectrum of COVID-19 neurology: clinical, radiological and laboratory findings. [cited 2020 Sep 28]; Available from: https://academic.oup.com/brain/advance-article/doi/10.1093/brain/awaa240/5868408

10. Puntmann VO, Carerj ML, Wieters I, Fahim M, Arendt C, Hoffmann J, et al. Outcomes of Cardiovascular Magnetic Resonance Imaging in Patients Recently Recovered from Coronavirus Disease 2019 (COVID-19). JAMA Cardiol [Internet]. 2020 [cited 2020 Sep 28]; Available from: https://jamanetwork.com/

11. Davies M-A. HIV and risk of COVID-19 death: a population cohort study from the Western Cape Province, South Africa. medRxiv Prepr Serv Heal Sci [Internet]. 2020 Jul 3 [cited 2020 Aug 19];2020.07.02.20145185. Available from: http://www.ncbi.nlm.nih.gov/pubmed/32637972

12. World Health Organisation. A Coordinated Global Research Roadmap: 2019 Novel Coronavirus. 2020.

13. African Academy of Sciences. Research and Development goals for COVID-19 in Africa The African Academy of Sciences Priority Setting Exercise [Internet]. Available from: https://www.aasciences.africa/sites/default/files/2020-04/Research.andDevelopmentGoalsforCOVID-19inAfrica.pdf

14. Norton A, De La Horra Gozalo A, Feune de Colombi N, Alobo M, Mutheu Asego J, Al-Rawni Z, et al. The remaining unknowns: a mixed methods study of the current and global health research priorities for COVID-19. BMJ Glob Heal [Internet]. 2020 Jul 29 [cited 2020 Aug 18];5(7):e003306. Available from: http://gh.bmj.com/lookup/doi/10.1136/bmjgh-2020-003306

15. Norton A, Mphahlele J, Yazdanpanah Y, Piot P, Tufet Bayona M. Strengthening the global effort on COVID-19 research. 2020; Available from: http://ees.elsevier.com/thelancet/www.thelancet.com

16. UKCDR & GloPID-R. COVID-19 Research Project Tracker [Internet]. 2020 [cited 2020 Jul 30]. Available from: https://www.ukcdr.org.uk/funding-landscape/covid-19-research-project-tracker/

17. Norton A, Bucher A, Antonio E, Advani N, Grund H, Mburu S, et al. Baseline results of a living systematic review for COVID-19 funded research projects. Wellcome Open Res [Internet]. 2020 Sep 8 [cited 2020 Sep 11];5:209. Available from: https://wellcomeopenresearch.org/articles/5-209/v1

18. COVID-19 R&D tracker – Policy Cures Research [Internet]. 2020 [cited 2020 Sep 1]. Available from: https://www.policycuresresearch.org/covid-19-r-d-tracker

19. Fitchett JRA, Lichtman A, Soyode DT, Low A, de Onis JV, Head MG, et al. Ebola research funding: A systematic analysis, 1997-2015. J Glob Health [Internet]. 2016 [cited 2020 Jul 19];6(2). Available from: /pmc/articles/PMC5112007/?report=abstract

20. NIH establishes Centers for Research in Emerging Infectious Diseases | National Institutes of Health (NIH) [Internet]. 2020 [cited 2020 Sep 1]. Available from: https://www.nih.gov/news-events/news-releases/nih-establishes-centers-research-emerging-infectious-diseases

21. World Health Organization (WHO). ACCESS TO COVID-19 TOOLS (ACT) ACCELERATOR COMMITMENT and CALL TO ACTION Our Vision and Mission. World Health Organization. 2020. p. 2020.

22. Götzinger F, Santiago-García B, Noguera-Julián A, Lanaspa M, Lancella L, Calò Carducci FI, et al. COVID-19 in children and adolescents in Europe: a multinational, multicentre cohort study. Lancet Child Adolesc Heal [Internet]. 2020 Jun [cited 2020 Jul 21];0(0). Available from: www.thelancet.com/child-adolescentPublishedonline

23. Ecdc. COVID-19 and schools transmission. 2020.

24. Grosse SD, Odame I, Atrash HK, Amendah DD, Piel FB, Williams TN. Sickle cell disease in Africa: A neglected cause of early childhood mortality. Vol. 41, American Journal of Preventive Medicine. Elsevier Inc.; 2011. p. S398–405.

25. UNICEF. Children, HIV and AIDS Global and regional snapshots [Internet]. 2019 [cited 2020 Aug 23]. Available from: https://data.unicef.org/resources/children-hiv-and-aids-global-and-regional-snapshots-2019/

26. The World staff. Zimbabwe health care workers strike over vital equipment [Internet]. 2020 [cited 2020 Aug 28]. Available from: https://www.pri.org/stories/2020-03-27/coronavirus-grows-zimbawe-health-care-workers-strike-over-vital-equipment

27. Reuters. Dozens of Kenyan doctors strike over lack of PPE, delayed pay [Internet]. 2020 [cited 2020 Aug 28]. Available from: https://www.reuters.com/article/us-health-coronavirus-kenya/dozens-of-kenyan-doctors-strike-over-lack-of-ppe-delayed-pay-idUSKCN25E20Q

28. Reuters. Nigerian doctors strike for better benefits during coronavirus crisis - Reuters [Internet]. 2020 [cited 2020 Aug 28]. Available from: https://uk.reuters.com/article/uk-health-coronavirus-nigeria-healthcare/nigerian-doctors-strike-for-better-benefits-during-coronavirus-crisis-idUKKBN23M1BM

29. Vivanti AJ, Vauloup-Fellous C, Prevot S, Zupan V, Suffee C, Do Cao J, et al. Transplacental transmission of SARS-CoV-2 infection. Nat Commun [Internet]. 2020 Jul 14 [cited 2020 Jul 23];11(1):3572. Available from: http://www.ncbi.nlm.nih.gov/pubmed/32665677

30. Karimi-Zarchi M, Neamatzadeh H, Dastgheib SA, Abbasi H, Mirjalili SR, Behforouz A, et al. Vertical Transmission of Coronavirus Disease 19 (COVID-19) from Infected Pregnant Mothers to Neonates: A Review [Internet]. Vol. 39, Fetal and Pediatric Pathology. Taylor and Francis Ltd; 2020 [cited 2020 Jul 23]. p. 246–50. Available from: /pmc/articles/PMC7157948/?report=abstract

31. World Health Organization (WHO). Global Breastfeeding Scorecard, 2017 Tracking Progress for Breastfeeding Policies and Programmes [Internet]. 2017. Available from: https://www.who.int/nutrition/publications/infantfeeding/global-bf-scorecard-2017.pdf?ua=1#:∼:text=The overall rate of exclusive,months of age is 40%25.&text=Nearly 40%;25 of the countries,countries have such high rates.

32. WHO Scientific Brief. Breastfeeding and COVID-19 [Internet]. 2020 [cited 2020 Jul 21]. Available from: https://www.who.int/news-room/commentaries/detail/breastfeeding-and-covid-19

33. Ghebreyesus TA. Opening remarks at the media briefing on COVID-19 [Internet]. World Health Organization. 2020 [cited 2020 Aug 23]. Available from: https://www.who.int/dg/speeches/detail/who-director-general-s-opening-remarks-at-the-media-briefing-on-covid-19---18-august-2020

34. Global Effort on COVID-19 (GECO) Health Research - Call Specification [Internet]. [cited 2020 Jul 23]. Available from: https://www.ukri.org/funding/funding-opportunities/global-effort-on-covid-19-geco-health-research/

35. Maher D, Aseffa A, Kay S, Tufet Bayona M. External funding to strengthen capacity for research in low-income and middle-income countries: exigence, excellence and equity. BMJ Glob Heal [Internet]. 2020 [cited 2020 Sep 30];5:2212. Available from: https://www.

36. Kilmarx PH, Maitin T, Adam T, Akuffo H, Aslanyan G, Cheetham M, et al. A Mechanism for Reviewing Investments in Health Research Capacity Strengthening in Low- and Middle-Income Countries. Ann Glob Heal [Internet]. 2020 Aug 3 [cited 2020 Sep 30];86(1):1–4. Available from: https://annalsofglobalhealth.org/articles/10.5334/aogh.2941/

